# Novel Genomic Risk Loci shared between Juvenile Idiopathic Arthritis and other HLA-associated Autoimmune Diseases

**DOI:** 10.64898/2026.01.21.26344521

**Authors:** Hamid Khoshfekr Rudsari, Piotr Jaholkowski, Srijana Bastakoti, Vera Fominykh, Kristine Løkås Haftorn, Alexey A. Shadrin, Benedicte A. Lie, Ole A. Andreassen, Helga Sanner

**Affiliations:** Department of Rheumatology, Oslo University Hospital, Oslo, Norway; Center for Precision Psychiatry, Division of Mental Health and Addiction, Oslo University Hospital, Oslo, Norway; Institute of Clinical Medicine, University of Oslo, Oslo, Norway; KG Jebsen Centre for Neurodevelopmental Disorders, University of Oslo, Oslo, Norway; Department of Medical Genetics, University of Oslo and Oslo University Hospital, Oslo, Norway; Department of Immunology, University of Oslo and Oslo University Hospital, Oslo, Norway

**Keywords:** Juvenile idiopathic arthritis, Autoimmune disease, Conjunctional false discovery rate, Genetic overlap, GWAS

## Abstract

**Objectives:** To investigate genetic architecture and identify novel risk loci shared between juvenile idiopathic arthritis (JIA) and other human leukocyte antigen (HLA)-associated autoimmune diseases (AIDs) by leveraging genome-wide association studies (GWAS) data.

**Methods:** We analyzed GWAS summary statistics from over two million participants (123,997 cases and 1,843,249 controls) of European ancestry across multiple AIDs including JIA, autoimmune thyroid disease (AITD), celiac disease, inflammatory bowel disease (IBD), multiple sclerosis (MS), psoriasis (Ps), rheumatoid arthritis (RA), systemic lupus erythematosus (SLE), and type 1 diabetes (T1D). The conjunctional false discovery rate (conjFDR) method was employed to identify shared genetic loci, followed by functional annotation and pathway analysis.

**Results:** We identified 46 novel loci shared between JIA and various AIDs: 17 with AITD, 16 with RA, 13 with IBD, five each with MS and Ps, four with SLE, and seven with T1D. Notable shared risk genes included *BACH2, UBASH3A, IRF4, IL12A-AS1, ETS1, PSMD14, FOXK1, NEK6, SP140, TNIP1, VAV3, CYP20A1, DNMT3A, LAX1, CASC15, ZC3H12C, KALRN, TBC1D1, PIK3CB, ANXA6*, and *HIF1A*. Functional annotation revealed 170 nonsynonymous exonic variants and 478 potentially deleterious variants (CADD score *>* 12.37). Gene Ontology analysis consistently highlighted enrichment of T cell-related processes and immune system regulation pathways.

**Conclusion:** This study expands the understanding of genetic architecture in JIA by identifying novel risk loci shared with other AIDs. The extensive genetic overlap and shared biological pathways, particularly in T cell-mediated immunity, suggest common pathogenic mechanisms and potential therapeutic targets across multiple autoimmune conditions.

## 1. Introduction

Juvenile idiopathic arthritis (JIA) is the most common chronic inflammatory rheumatic disease in children, defined as arthritis of unknown cause beginning before age 16 and lasting at least six weeks [1, 2]. It comprises seven clinically distinct subtypes according to the International League of Associations for Rheumatology (ILAR) classification [2]. Despite its clinical importance, JIA’s relatively low prevalence poses substantial challenges for genetic studies, with limited sample sizes constraining the statistical power of traditional genomewide association studies (GWAS) approaches. The largest to-date JIA GWAS included 3,305 patients and 9,196 controls, identifying five genetic loci [3]. Using fine-mapping and integrative analysis, researchers pinpointed potential causal single nucleotide polymorphisms (SNPs) and highlighted transcription factors *RELA* and *EBF1* as contributors to disease risk, while identifying *IL6ST* as a potential drug target linking risk loci to specific transcriptional regulators and cytokine pathways [3].

Like other autoimmune diseases (AIDs), JIA has well-established genetic associations with the human leukocyte antigen (HLA) region [4, 5], which plays a crucial role in immune response and pathogenesis. This genetic architecture is shared broadly across AIDs—a group of over 80 chronic illnesses marked by immune system dysfunction, leading to loss of self-tolerance and chronic inflammation [6]. Although different AIDs may involve distinct organ systems, they share a complex genetic basis [7]. GWAS have revealed that the HLA region is strongly associated with most AIDs including rheumatoid arthritis (RA), systemic lupus erythematosus (SLE), type 1 diabetes (T1D), multiple sclerosis (MS), celiac disease, psoriasis (Ps), autoimmune thyroid disease (AITD), and inflammatory bowel disease (IBD), indicating extensive genetic overlap [8, 9]. Additionally, non-HLA genes central to autoimmune inflammation such as *IL23R, TNFAIP3*, and *IL2RA* are implicated in multiple AIDs, supporting shared genetic pathways [10].

Studies have demonstrated significant overlaps in genetic risk factors between JIA and other AIDs, particularly RA, with shared susceptibility loci such as *STAT4* (mediating signaling downstream of *IL-12* and *IL-23*, and enhancing differentiation of Th1 and Th17 cells), *TRAF1/C5* (involved in TNF receptor signaling and complement activation), and *CD247* (crucial for T-cell receptor signal transduction) [11, 12]. These findings highlight extensive genetic overlap and shared molecular pathways underlying autoimmunity, offering avenues for improved disease classification and treatment. Researchers have successfully identified JIA susceptibility genes such as *PTPN22* [13] and *IL2RA* [14] using multi-AID approaches that examine shared genetic architecture across related autoimmune conditions.

Given JIA’s low prevalence and consequently limited sample sizes for genetic studies [3], traditional GWAS approaches face substantial power constraints. A cross-disease framework that leverages shared genetic architecture with other AIDs provides a powerful complementary strategy to identify additional risk loci. This approach not only enhances statistical power for locus discovery but also offers insights into shared pathogenic mechanisms that may inform therapeutic repositioning strategies across AIDs. In this study, we enhance understanding of genetic relationships between JIA and eight major AIDs using the conjunctional false discovery rate (conjFDR) approach. This methodology boosts discovery by leveraging overlapping GWAS associations [15, 16]. This strategy not only expands understanding of JIA’s genetic architecture but also identifies novel risk loci shared across multiple autoimmune conditions, paving the way for more effective diagnostics and therapeutics.

## 2. Methods

### 2.1. GWAS Description

We obtained summary statistics from available GWASs with the largest sample sizes for each AID (Table 1). We excluded overlapping samples to avoid biases. In total, we analyzed GWAS data from around 2 million participants (123,997 cases and 1,843,249 controls).

**Table 1:** Summary data of all GWASs used in this paper.

| Disease | Sample size, n | SNPs, n | Source |
| --- | --- | --- | --- |
| JIA | 3,305 cases, 9,196 controls | 7461261 | López-Isac et al. [3] |
| AITD | 30,234 cases, 725,172 controls | 34819036 | Saevarsdottir et al. [23] |
| Celiac disease | 1,036 cases, 455,312 controls | 11842647 | Jiang et al. [18] |
| IBD | 12,160 cases, 13,145 controls | 9928119 | Lange et al. [24] |
| MS | 14,802 cases, 26,703 controls | 8458297 | Patsopoulos et al. [25] |
| Ps | 15,967 cases, 28,194 controls | 9467953 | Stuart et al. [19] |
| RA | 22,350 cases, 74,823 controls | 12661745 | Ishigaki et al. [21] |
| SLE | 5,201 cases, 9,066 controls | 644674 | Bentham et al. [20] |
| T1D | 18,942 cases, 501,638 controls | 62116689 | Chiou et al. [22] |

GWAS summary statistics were downloaded from the NHGRI-EBI GWAS Catalog [17] for studies GCST90010715 [3] (JIA), GCST90044158 [18] (celiac disease), GCST90019016 [19] (Ps), GCST003156 [20] (SLE), GCST90132222 [21] (RA), and GCST90014023 [22] (T1D). AITD and IBD statistics were obtained from meta-analyses [23, 24], and MS from the International Multiple Sclerosis Genetics Consortium [25]. All participants were of European ancestry.

### 2.2. Statistical Analyses

#### 2.2.1. Conjunctional false discovery rate analysis

We utilized the conjFDR method to enhance identification of genomic loci jointly associated with JIA and other AIDs. This approach builds on an empirical Bayes mixture model [26], offering higher replication rates and previously applied successfully to various phenotypes [27, 28]. The conditional false discovery rate (condFDR) is the posterior probability that a SNP is null for the primary trait given that *P*-values for both traits are as small or smaller than observed values, while conjFDR identifies shared genetic loci as the posterior probability that a SNP is null for either one or both traits [29]. We adopted conjFDR and condFDR thresholds of 0.05 and 0.01 respectively [30]. To prevent bias from complex regional linkage disequilibrium (LD), we excluded SNPs in the extended MHC region (chr6:25119106-33854733) and 8p23.1 inversion before applying the FDR model [27, 5, 31], and visualized cross-trait enrichment using conditional Q-Q plots.

### 2.3. Functional Analyses

#### 2.3.1. Genomic loci definition

We followed the FUMA protocol [32] to identify independent significant SNPs, defined as SNPs with a conjFDR < 0.05 and independent of each other at *r*^2^ < 0.6. SNPs with conjFDR < 0.10 that were in LD (*r*^2^ *>* 0.6) with any of the independent significant SNPs were defined as candidate SNPs. For each independent significant SNP, locus boundaries were defined as the minimum and maximum coordinates of the corresponding candidate SNPs. Loci within 250 kilobases (kb) of each other were merged into a single locus. Lead SNPs were defined as independent significant SNPs that were independent of each other at *r*^2^ < 0.1. For each locus, the locus lead variant was defined as the lead SNP with the lowest conjFDR. LD *r*^2^ values were calculated using European-ancestry samples from the 1000 Genomes Project Phase 3 reference panel.

We assessed directional effects of shared loci by comparing z-scores and odds ratios. Novel loci were considered novel if not located within ±1 Mb of previously reported loci [3, 33, 34, 35, 4, 36, 37, 38, 39]. When reporting novel loci, overlapping lead SNPs are reported as a single locus. We may report different lead SNPs for each novel locus shared between JIA and AIDs but they correspond to the same locus.

#### 2.3.2. Functional annotation

SNPs were functionally annotated using combined annotation dependent depletion (CADD) scores [40], regulomeDB scores, and chromatin states. CADD scores predict potential harmful impact on proteins, regulomeDB scores estimate regulatory function likelihood, and chromatin states indicate potential transcriptional effects. Candidate SNPs were linked to potential causal genes through positional mapping, expression quantitative trait locus (eQTL) mapping, and chromatin interaction mapping. Gene expression and gene-set analysis were conducted using FUMA and Genotype-Tissue Expression (GTEx) data.

## 3. Results

### 3.1. Cross-trait enrichment

Figure 1 shows conditional Q-Q plots demonstrating significant enrichment of SNP associations with JIA as SNP association levels increase across all AIDs, indicating substantial polygenic overlap. The initial leftward deviation from the dashed line indicated a higher proportion of genuine associations for a given nominal JIA *P*-value. Further leftward shifts at decreasing nominal *P*-value thresholds showed that the proportion of genuine effects in JIA increases as association strength with each AID increases. As shown in Figure 1, we also observed enrichment of SNP associations with all considered AIDs conditional on JIA.

**Figure 1:**
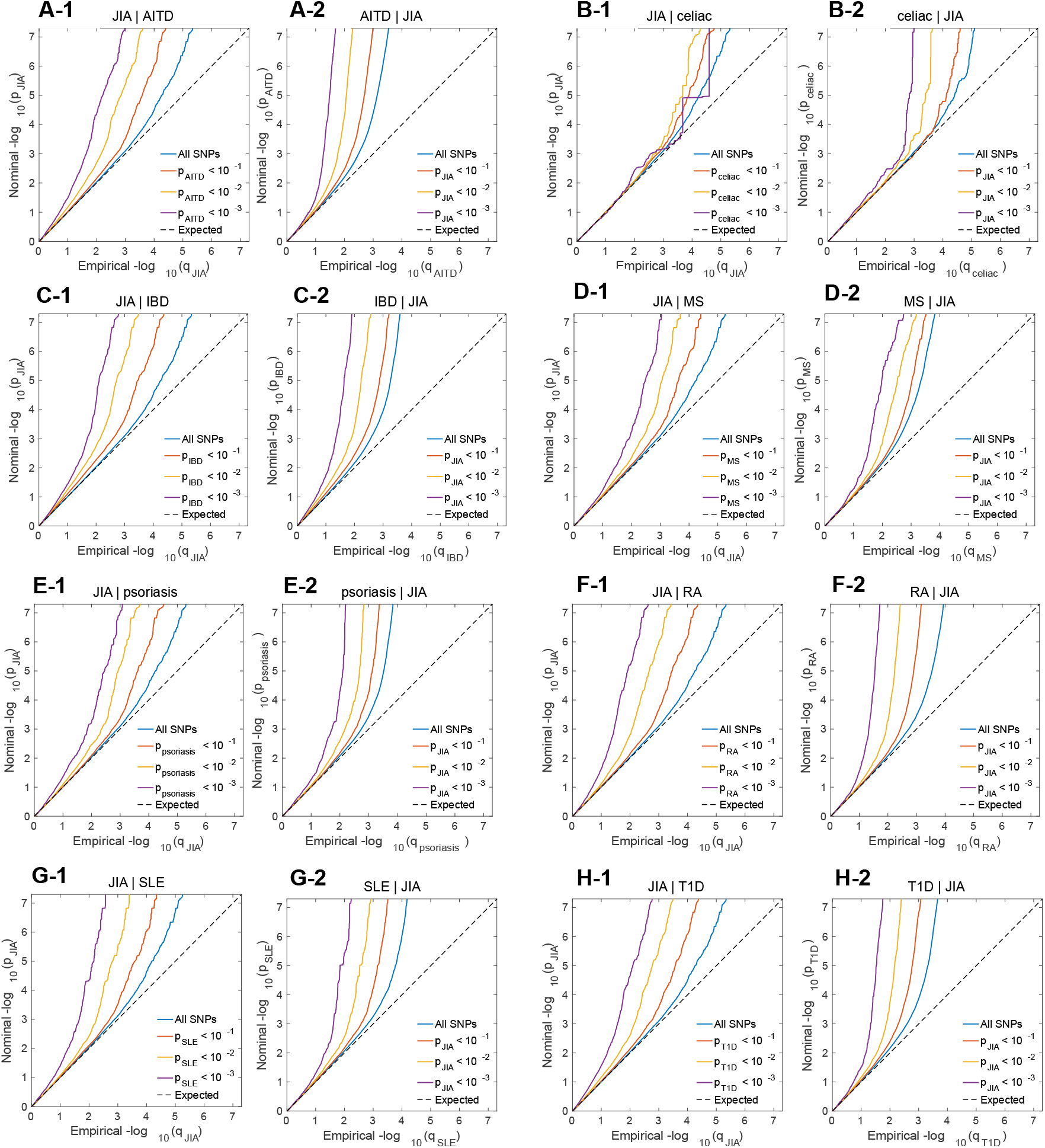
Cross-trait enrichment between JIA and AIDs. Quantile-quantile (Q-Q) plots show SNP enrichment for JIA conditional on SNP associations with AIDs and reverse conditional Q-Q plots. (A) Autoimmune thyroid disease (AITD), (B) celiac disease, (C) inflammatory bowel disease (IBD), (D) multiple sclerosis (MS), (E) psoriasis, (F) rheumatoid arthritis (RA), (G) systemic lupus erythematosus (SLE), and (H) type-1 diabetes (T1D). Conditional Q-Q plots of nominal versus empirical −log10 *P*-values (corrected for inflation) in JIA below *P* <*×*5 10^*−*8^ as a function of significance with AIDs at *P* < 0.10, *P* < 0.01 and *P* < 0.001. Blue lines indicate all SNPs. Dashed lines indicate the null hypothesis.

### 3.2. Shared loci between JIA and the AIDs

We utilized bi-directional cross-trait enrichment to enhance statistical power for discovering shared loci. At conjFDR < 0.05, we identified 78 loci shared between JIA and AITD, one with celiac disease, 68 with IBD, 28 with MS, 27 with Ps, 61 with RA, 19 with SLE, and 54 with T1D (Figure 2 and Supplementary Tables 1-8). Overall, we identified 46 shared loci which are novel for JIA given in Table 2.

**Table 2:**
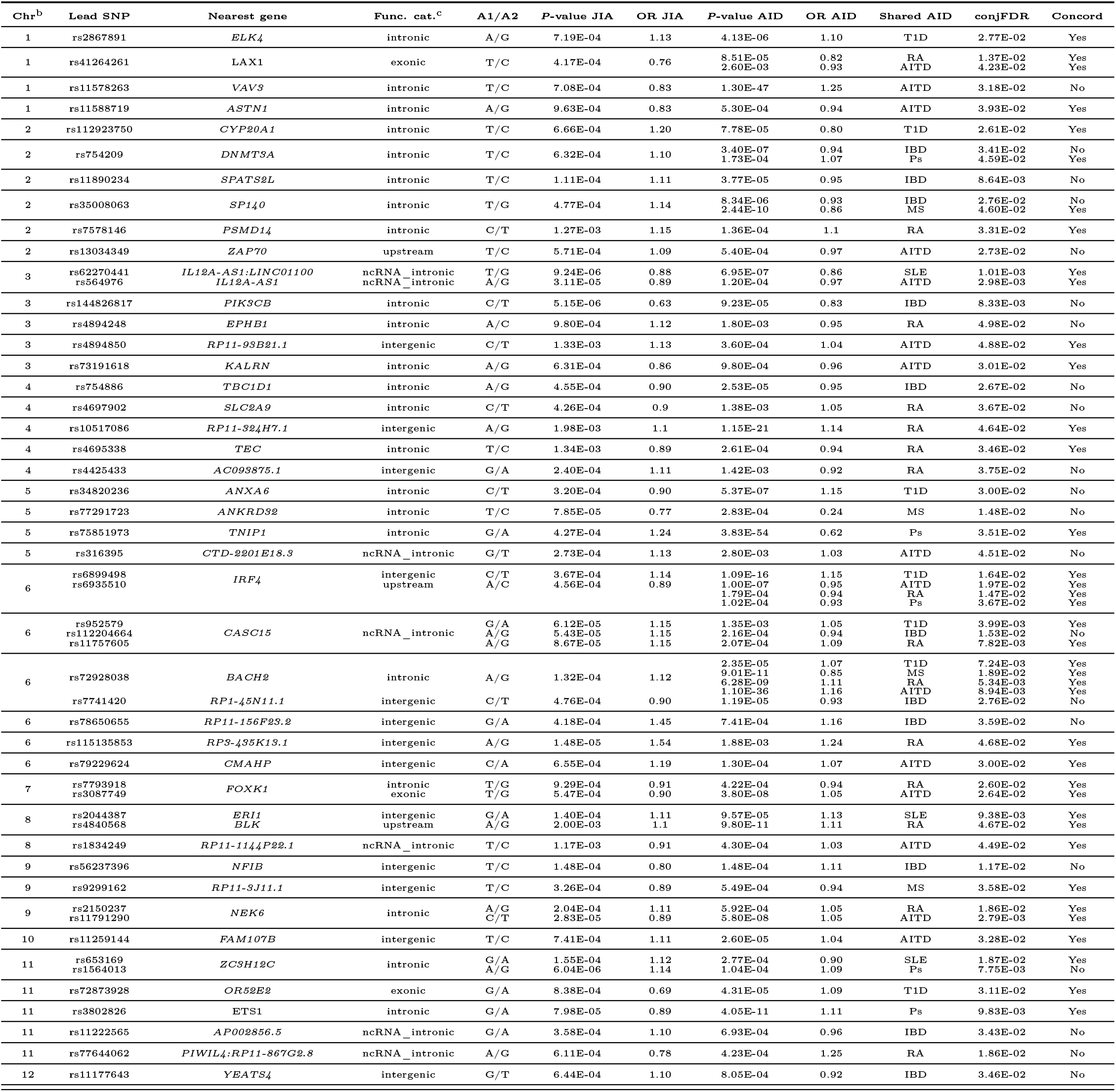
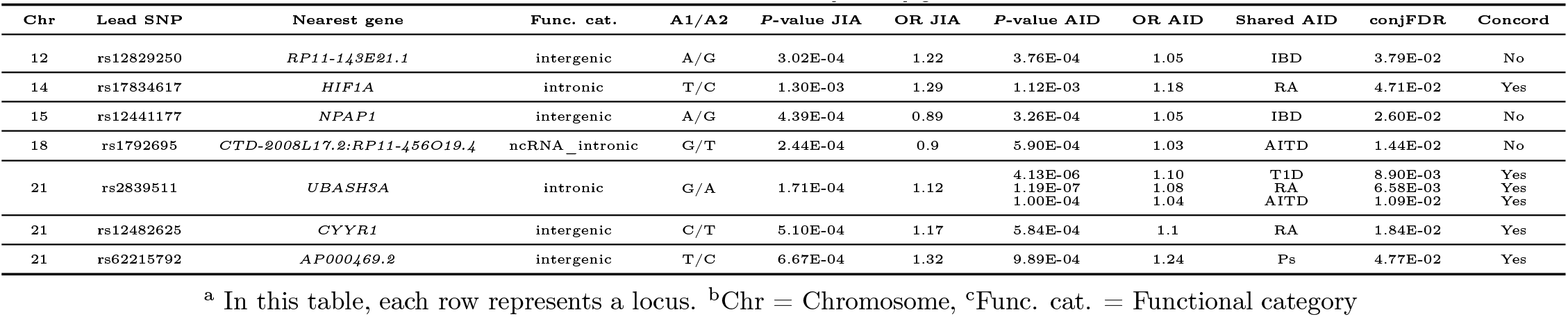
Novel JIA genomic loci also associated with other AIDs at conjFDR < 0.05^a^.

**Figure 2:**
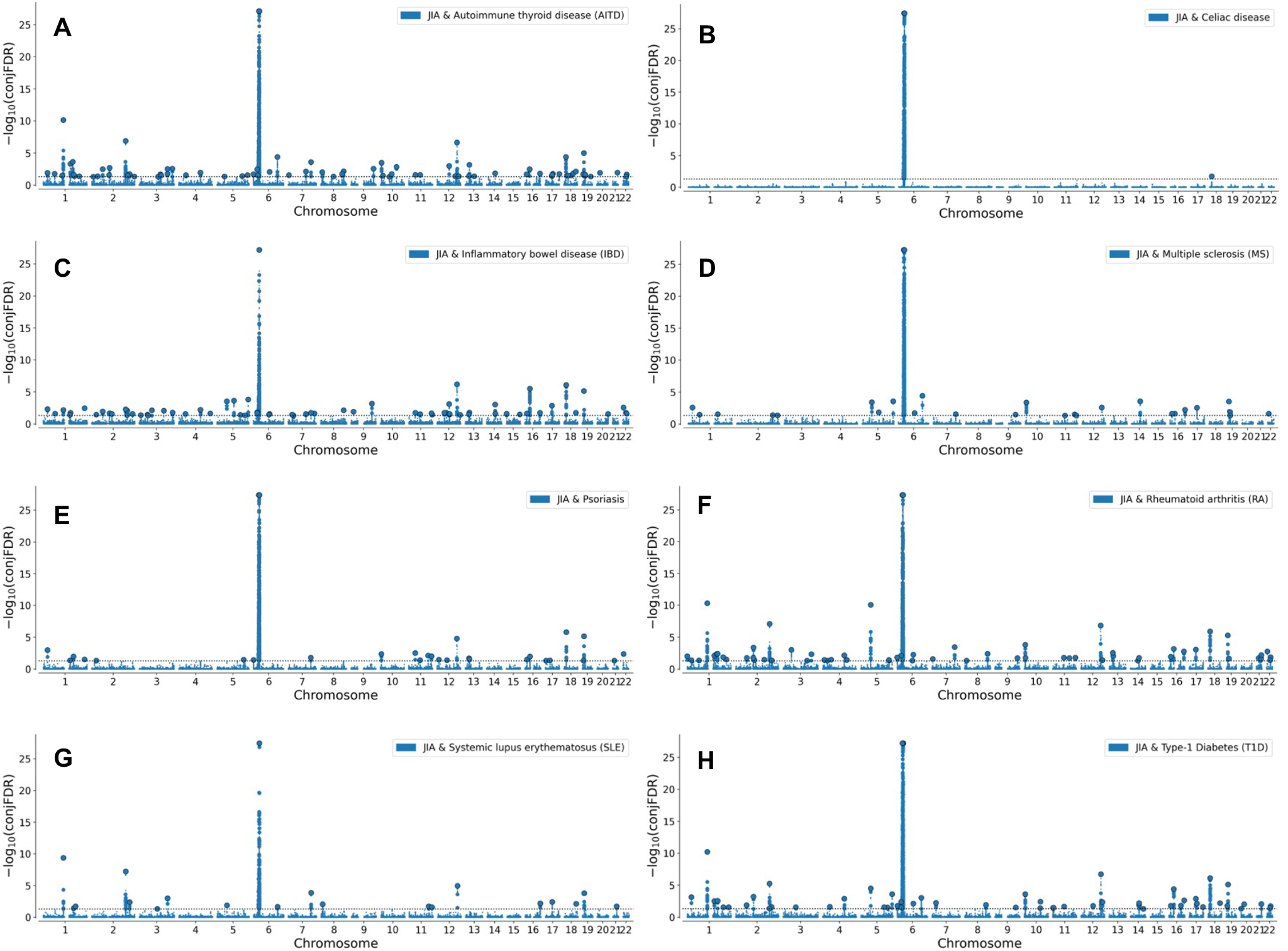
Shared loci between JIA and AIDs at conjFDR < 0.05. Common genetic variants jointly associated with JIA and (A) AITD, (B) celiac disease, (C) IBD, (D) MS, (E) psoriasis, (F) RA, (G) SLE, and (H) T1D. Manhattan plots showing −log10 transformed conjFDR values for each SNP on the y-axis and chromosomal positions on the x-axis. Dotted horizontal lines represent the threshold for significant shared associations [conjFDR < 0.05, i.e. −log10(conjFDR) > 1.3]. Independent lead SNPs are circled in black. Significant shared signals in the MHC region are represented by one lead SNP only.

Novel JIA loci were distributed across multiple chromosomes with distinct sharing patterns. We found 17 novel loci shared between AITD across ten chromosomes, 16 loci shared with RA across ten chromosomes, and 13 loci shared with IBD across eight chromosomes. For MS and Ps, we identified five novel loci each across five and four chromosomes, respectively. SLE shared four novel loci with JIA, while T1D shared seven. Several loci showed notable pleiotropy. For example, *BACH2* on chromosome 6 was associated with T1D, RA, AITD, and MS, while *UBASH3A* on chromosome 21 showed associations with T1D, RA, and AITD.

We evaluated effect directions of lead SNPs for all shared loci (Supplementary Tables 1-8, Table 3). High concordance of effect directions was observed between JIA and several AIDs, suggesting shared underlying immunopathology. According to Table 3, more than 80% of shared loci between JIA and each of the studied AIDs had concordant allelic effect directions, except for IBD, which exhibited predominantly (85%) opposite effect directions.

**Table 3:** Direction of allelic effects for shared genetic loci between JIA and other AIDs.

| AID | Total shared loci | Concordant direction (%) | Opposite direction (%) |
| --- | --- | --- | --- |
| AITD | 78 | 63 (81%) | 15 (19%) |
| Celiac disease | 1 | 1 (100%) | 0 (0%) |
| IBD | 68 | 10 (15%) | 58 (85%) |
| MS | 28 | 23 (82%) | 5 (18%) |
| Psoriasis | 27 | 24 (89%) | 3 (11%) |
| RA | 61 | 51 (84%) | 10 (16%) |
| SLE | 19 | 17 (89%) | 2 (11%) |
| T1D | 54 | 47 (87%) | 7 (13%) |

### 3.3. Functional annotation

Functional annotation revealed that candidate SNPs were predominantly located in intergenic and intronic regions, with 170 nonsynonymous exonic variants identified across JIAAID shared loci (Supplementary Tables 9-17). The most broadly shared variants implicated key immune regulatory genes including *SH2B3, TYK2*, and *PDE4A* (shared across JIA and eight AIDs), while disease-specific patterns emerged for genes such as *PTPN22, CD226*, and *RUNX3* (Supplementary Tables 9-17).

Among shared loci, 478 candidate SNPs exhibited CADD scores *>* 12.37, indicating potential deleteriousness, with the highest numbers observed in loci shared between JIA and AITD (46 SNPs) and JIA and RA (45 SNPs) comparisons. Three-way gene mapping identified 336 distinct loci linked to 1,300 genes (Supplementary Tables 18-25).

Gene Ontology analysis revealed consistent enrichment of immune-related pathways across loci shared between JIA and AIDs as shown in Figure 3. The most significant biological processes centered on T cell activation and differentiation, cytokine-mediated signaling, and immune system regulation (Supplementary Tables 26-31). Notably, JIA-AITD shared loci were enriched for phosphatase binding and interleukin-2 signaling, while JIA-IBD pairs showed strong enrichment for cytokine response and bacterial defense pathways (Supplementary Tables 26-31). These findings highlight conserved immune regulatory mechanisms underlying JIA’s overlap with multiple autoimmune conditions. The gene-set analyses for the other groups of shared loci lacked sufficient power.

**Figure 3:**
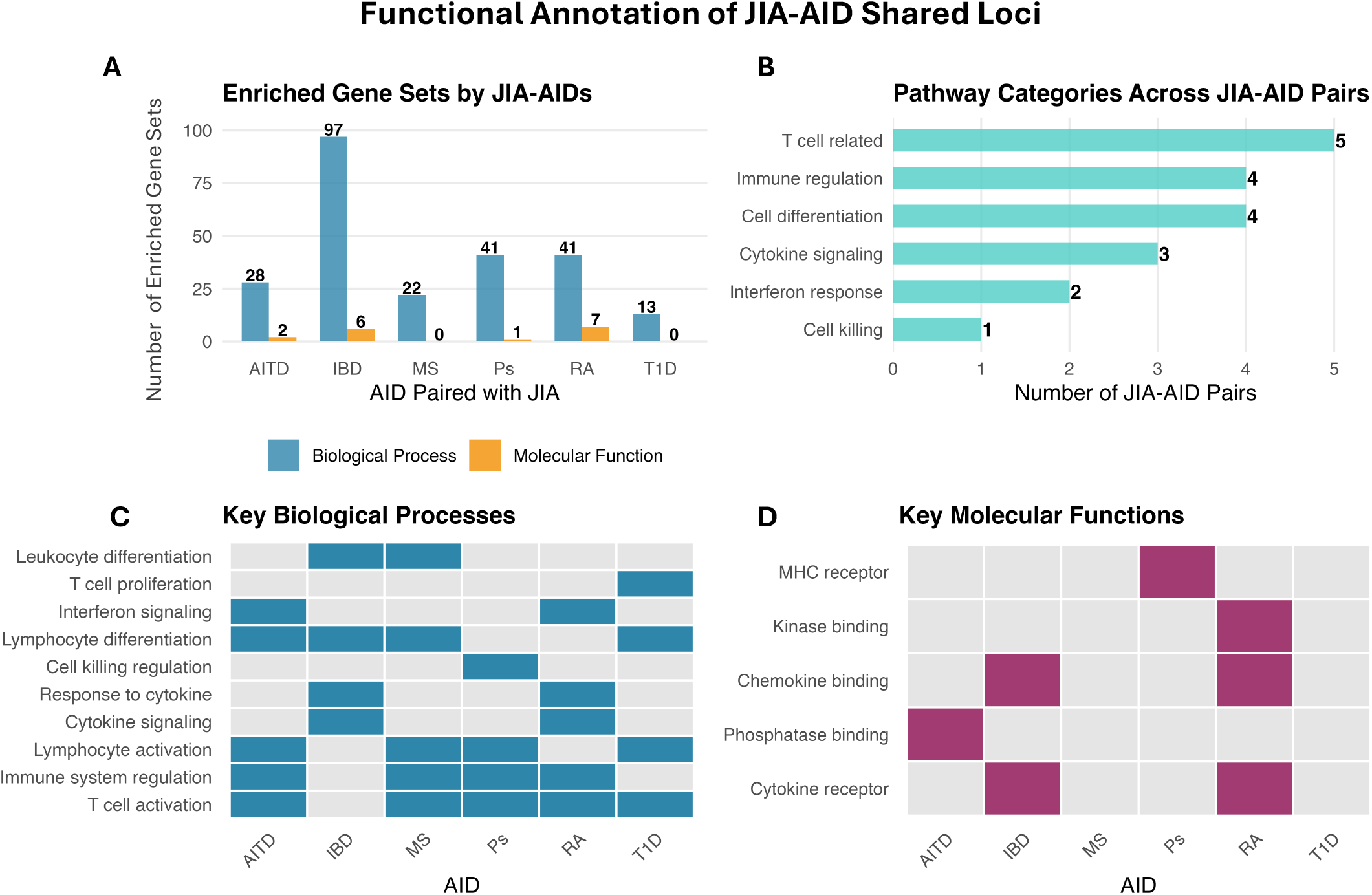
Functional enrichment analysis of JIA-AIDs shared loci. Shared genetic loci between JIA and seven AIDs show significant enrichment for immune regulatory pathways. (A) Number of enriched gene sets per disease pair. (B) Pathway category distribution. (C,D) Heatmaps showing enriched biological processes and molecular functions, with T cell activation and cytokine signaling as predominant themes.

## 4. Discussion

This study expands understanding of JIA’s genetic architecture through identification of 46 novel genomic risk loci shared between eight major AIDs. By employing conjFDR analysis across around two million participants, we demonstrate extensive genetic pleiotropy between JIA and other AIDs, with particularly strong overlap observed for AITD (17 novel loci), RA (16 loci), and IBD (13 loci). These findings enhance statistical power for genetic discovery in JIA and reveal common pathogenic mechanisms that may inform therapeutic strategies across multiple autoimmune conditions. Collectively, the findings reveal an extensive shared genetic architecture between JIA and multiple AIDs, converging on T cell-regulatory pathways, epigenetic control, and cytokine signaling.

### 4.1. Convergence on Core Immune Regulatory Pathways

Functional analyses reveal that shared genetic architecture between JIA and other AIDs converges on central immune regulatory mechanisms. Gene Ontology analysis consistently highlighted T cell-related processes, including T cell activation, differentiation, and cytokine-mediated signaling. This pattern suggests that dysregulation of T cell immunity represents a fundamental mechanism underlying autoimmune susceptibility across disease phenotypes [41].

Several genes exemplify this convergence. *BACH2*, identified on chromosome 6 and shared across T1D, RA, MS, and AITD, encodes a transcription factor critical for multiple immune lineages. Its role in promoting antibody class switching in germinal center B cells while suppressing plasma cell differentiation [42], combined with its guidance of Th cell differentiation [43], positions it as a master regulator of adaptive immunity. Similarly, *UBASH3A* on chromosome 21, shared among T1D, RA, and AITD, functions as a key negative regulator of both TCR and NF-*κ*B signaling [44], with disease-associated variants correlating with increased expression and altered T cell responses [45].

### 4.2. Epigenetic Regulation and Transcriptional Networks

Beyond classical immune signaling molecules, our analysis identified genes governing epigenetic modification as shared risk factors. *DNMT3A* (chromosome 2), associated with both IBD and psoriasis, regulates DNA methylation [46, 47, 48]. *SP140*, an immune-restricted epigenetic reader, functions as a critical repressor of inflammation through NF-*κ*B-mediated mechanisms [49]. Long non-coding RNAs also emerged as shared risk factors, including *CASC15* and *IL12A-AS1* on chromosome 3. *IL12A-AS1* contains regulatory elements in transcription factor binding sites of immune cells, particularly monocytes and macrophages [50].

The transcription factors *IRF4, ETS1*, and *FOXK1* represent key nodes in shared genetic networks. *IRF4* ‘s essential role in Th cell, Treg, B cell, and dendritic cell development [51] positions it as a critical determinant of immune homeostasis. *ETS1*, showing associations with SLE, RA, IBD, and psoriasis, controls B cell autoimmune responses, with deficiency leading to hyperresponsive B cells and autoantibody production [52]. Recent identification of *FOXK1* in AITD studies [53] and its role in the HDAC3-FOXK1-interferon axis in RA [54] suggest conserved transcriptional programs across multiple autoimmune conditions.

Functional annotation identified 170 nonsynonymous exonic variants and 478 potentially deleterious variants (CADD score > 12.37) across shared loci. Genes with the broadest sharing patterns—including *SH2B3, TYK2*, and *PDE4A*—represent established therapeutic targets, supporting potential for drug repurposing across AIDs.

### 4.3. Clinical Implications and Therapeutic Opportunities

The extensive genetic overlap between JIA and adult AIDs provides molecular support for shared therapeutic approaches. Current successful translation of RA therapeutics to JIA management [55, 56] aligns with our identification of 16 shared genetic loci between these conditions. Beyond RA, the shared architecture with IBD (13 loci), MS (5 loci), and T1D (7 loci) suggests potential efficacy of biologics targeting shared pathways, such as IL-12/IL-23 inhibitors and JAK-STAT pathway modulators.

Direction of allelic effects provides insight into disease relationships. The high concordance of effect directions between JIA and AITD (63/78 loci), T1D (47/54 loci), and SLE (17/19 loci) suggests similar pathogenic mechanisms, whereas more discordant patterns with IBD (58/68 opposite directions) may reflect distinct tissue-specific immune processes despite shared genetic susceptibility.

### 4.4. Limitations and Future Directions

Several considerations contextualize our findings. First, exclusion of the extended MHC region, while methodologically necessary to avoid LD bias in conjFDR analysis, means our results do not capture the full spectrum of shared genetic architecture in this immunologically critical region. Second, the smaller JIA sample size relative to adult AID cohorts may limit detection of additional shared associations with smaller effect sizes. Third, our focus on common variants does not address potential contributions of rare variants to disease risk.

Functional validation of identified variants, particularly those with high deleteriousness scores and pleiotropic effects, represents a critical next step. Integration of genetic findings with single-cell transcriptomics and epigenomic profiling will elucidate cell-type-specific mechanisms. Development of polygenic risk scores incorporating shared genetic architecture may enable earlier disease prediction and personalized therapeutic approaches.

### 4.5. Conclusion

This comprehensive genetic analysis reveals that JIA shares substantial genetic architecture with multiple AIDs, with convergence on T cell-mediated immunity, epigenetic regulation, and inflammatory signaling pathways. Overall, the identification of 66 novel shared loci implicates core immune pathways and reveals targets with relevance for disease mechanisms, diagnostics, and therapeutic repurposing.

## Data Availability

All data produced are available online at NHGRI-EBI GWAS Catalog.

## Acknowledgments

This work is partially funded by Helse Sør-øst Rhf Grant number HSO - 2022 / 202307.

## Declaration of generative AI and AI-assisted technologies in the manuscript preparation process

The authors declare that generative AI was used in the creation of this manuscript. ChatGPT was solely used to improve the language clarity and precision, as English is not the primary language of the authors. It was not used for data analysis or any other purpose beyond improving language.

